# A community-based ANP-led integrated oncology care model for adults receiving oral anticancer medication-protocol for a pilot study

**DOI:** 10.1101/2022.02.23.22271044

**Authors:** J.P. Richmond, A. Johnston, P.J. Murphy, P. Gillespie, A.W. Murphy

## Abstract

Oncology has been undergoing a profound transition in the last ten years or more with the increased usage in oral anti-cancer medication (OAM). Approximately 25% of all anti-cancer medication is now designed for oral use and this is likely to increase prospectively. Oral anti-cancer medications have the potential to alleviate capacity issues in cancer treating units as patients receive their treatment at home. There remains however a requirement for safe and efficient assessment and monitoring but this does not necessarily require them to repeatedly attend a hospital day unit. Therefore the opportunity exists to transition this cohort to a community-based setting to be assessed by a specialist such as an Advanced Nurse Practitioner (ANP) in nurse-led clinics. Having an OAM assessment closer to their home would be more convenient to the patient. Furthermore, this could help alleviate hospital capacity issues which were brought into sharp focus with the onset of the COVID-19 pandemic and the use of nurse-led clinics are promoted in the aims of the current healthcare system reform process in Ireland.

Within the context of the Irish healthcare system reform and the COVID-19 pandemic this protocol will outline a collaboration between an Oncology Department in Letterkenny University Hospital in Ireland and the National University of Ireland, Galway aimed to develop and pilot a community-based Advanced Nurse Practitioner-led integrated oncology care model for adults receiving OAM. Phase 1 of this two-phase study commenced in September 2020 and comprised a scoping review, a benchmarking exercise and a qualitative analysis of relevant stakeholders. This protocol paper presents a pilot to be undertaken in phase 2 as OAM care is transitioned to an ANP-led community-based model, which is a radical shift for oncology care in Ireland. The pilot outlined will provide data that will identify potential refinements to the model and address specific uncertainties about a definitive trial.

## Introduction

The emergence of oral anti-cancer medication (OAM) has revolutionised oncology care, particularly within the last decade and these treatments are being approved at a record setting pace (Meier et al., 2018). Oral anticancer medications are a sub-set of systemic anti-cancer treatments (SACT) which are administered enterally with a narrow therapeutic window and a unique mechanism of action (National Cancer Control Programme (NCCP), 2018; National Cancer Institute, 2020). OAMs have the same benefits and risks as SACT given intravenously in terms of positive disease outcomes, treatment-related toxicities, and potential for serious medication errors leading to patient harm (NCCP, 2018). While these medications are convenient and often preferred or even requested by patients, it shifts the responsibility for medication management from the oncology healthcare professionals (HCPs) to the patient. Consequently, there are concerns regarding adherence and patients’ management of toxicities or adverse effects (Greer et al., 2016; Hammond et al., 2012; NCCP, 2018; Paolella et al., 2018; Wood, 2012).

Due to the associated safety challenges, ongoing specialised assessment and monitoring of patients receiving OAMs is essential; the organisation of this is usually not much different to parenteral anti-cancer treatment. In Ireland, historically the practice has been that the patient generally attends the Oncology or Haematology Day Unit for a dedicated health assessment, which includes review of recent laboratory tests and/or other investigative results (Hammond et al, 2012; Department of Health (DOH), 2017; NCCP, 2018). This practice, while necessary for the aforementioned reasons, when performed in the context of increasing incidence and treatment of cancer, has contributed to hospital overcrowding (Sung et al, 2021; DOH, 2017).

### The Impact of the COVID-19 Pandemic

Such pre-existing hospital capacity issues were exacerbated with the advent of the COVID-19 pandemic and the requirement for social distancing. Since March 2020, internationally, HCPs have been forced to reassess routine practices (Volger & Lightner, 2020) and to reduce unnecessary patient hospital visits (Cucinotta & Vanelli, 2020). This has been especially pertinent to individuals receiving cancer treatment due to potential immunosuppression and concomitant risk of infection (Leung et al, 2020).

At the Letterkenny University Hospital (LUH) Oncology Department (institution of authors JR, AJ, and MGK), the onset of the pandemic required the immediate transfer of the caseload of individuals receiving OAMs from the Oncology Day Unit to the Advanced Nurse Practitioner (ANP) Oncology Clinic for ongoing assessment. This was in order to maintain COVID restrictions and free up capacity for patients receiving intravenous SACT. This is an example of a successful nurse-led clinic. Nurse-led clinics have emerged internationally as an ideal means to achieve improved organisation and efficiency in health services (Randall et al., 2017, House of the Oireachtas, 2018; Torrens et al, 2020), with associated high levels of patient satisfaction (Molassiotis et al., 2020, Linedale et al., 2020), and potential cost-savings (Thompson & McNamara, 2021).

### The Shift to Primary Care

While there is a requirement for patients receiving OAMs to have on-going assessment and monitoring, this does not necessarily require them to repeatedly attend a hospital day unit. The opportunity exists to transition this cohort to a community-based setting.

Transitioning to a community-based setting would align with the Government of Ireland’s Sláintecare reform (House of the Oireachtas, 2018), which aims to shift healthcare from hospitals to the community. Indeed, internationally there is a transformative vision to shift patient care from acute to primary care, with a universal consensus regarding the crucial and central role for primary care (Calnan et al., 2006; Richmond et al., 2021).

### The Current Study

With this background of Irish health service reform and the COVID-19 pandemic, a collaboration between the LUH Oncology Department and the National University of Ireland, Galway (NUIG) aimed to develop and pilot a community-based ANP-led integrated oncology care model for adults receiving OAM.

Phase 1 of this two-phase study commenced in September 2020 and comprised of three elements: a scoping review, a benchmarking exercise and a qualitative analysis of relevant stakeholders. The scoping review aimed to determine current clinical management practices for the ongoing assessment and monitoring of patients receiving OAM (Richmond, 2021a). The authors reported that in the studies reviewed that there was a unanimous endorsement of a dedicated OAM clinic as a means to achieve improved care for this patient cohort. The scoping review also identified a range of best-practice recommendations for clinical practice, which were collated alongside existing national and international guidelines to develop a benchmarking tool. A member of the research team, who was not actively involved in the day-to-day OAM clinic, used this tool to retrospectively examine the standard of existing care of the ANP-led hospital–based OAM clinic. This benchmarking exercise demonstrated safe practice yet noted scope for improvement, especially with regard to documentation of patient education and standardisation of OAM prescription writing. The third piece of work in Phase 1 was a qualitative analysis of relevant stakeholders’ (nurses, doctors, pharmacists, general practitioners, and service users) perceptions of a community-based ANP-led integrated model for OAM care as well as identification of the infrastructural and cultural supports required for this to happen.

The research team presented the results from the three elements of Phase 1 to an advisory panel of local and national experts in October 2021. The outcome was an agreement that a pilot study should proceed to examine the feasibility of a community-based ANP-led integrated oncology care model for adults receiving OAMs. The results of Phase 1 have directly informed the development of the protocol for Phase 2, presented in the remainder of this article.

## Phase 2: Pilot Study Protocol

A pilot study is essential to assess the acceptability and feasibility of interventions and study protocols (Skivington et al, 2021), and can prevent the cost of a failed trial (Arain et al, 2010). Richards & Halberg (2015) identified that commonly researchers fail to adequately develop and pilot phases of trials. As the authors propose transforming OAM care to an ANP-led community-based model, which is a radical shift for oncology care in Ireland, a pilot will provide data that will identify potential refinements to the model and address specific uncertainties about a definitive trial. We use the framework of 14 questions identified by Shanyinde at al (2011) that should be asked and answered in a pilot trial. These are presented in Table 1.

**Table 1:**
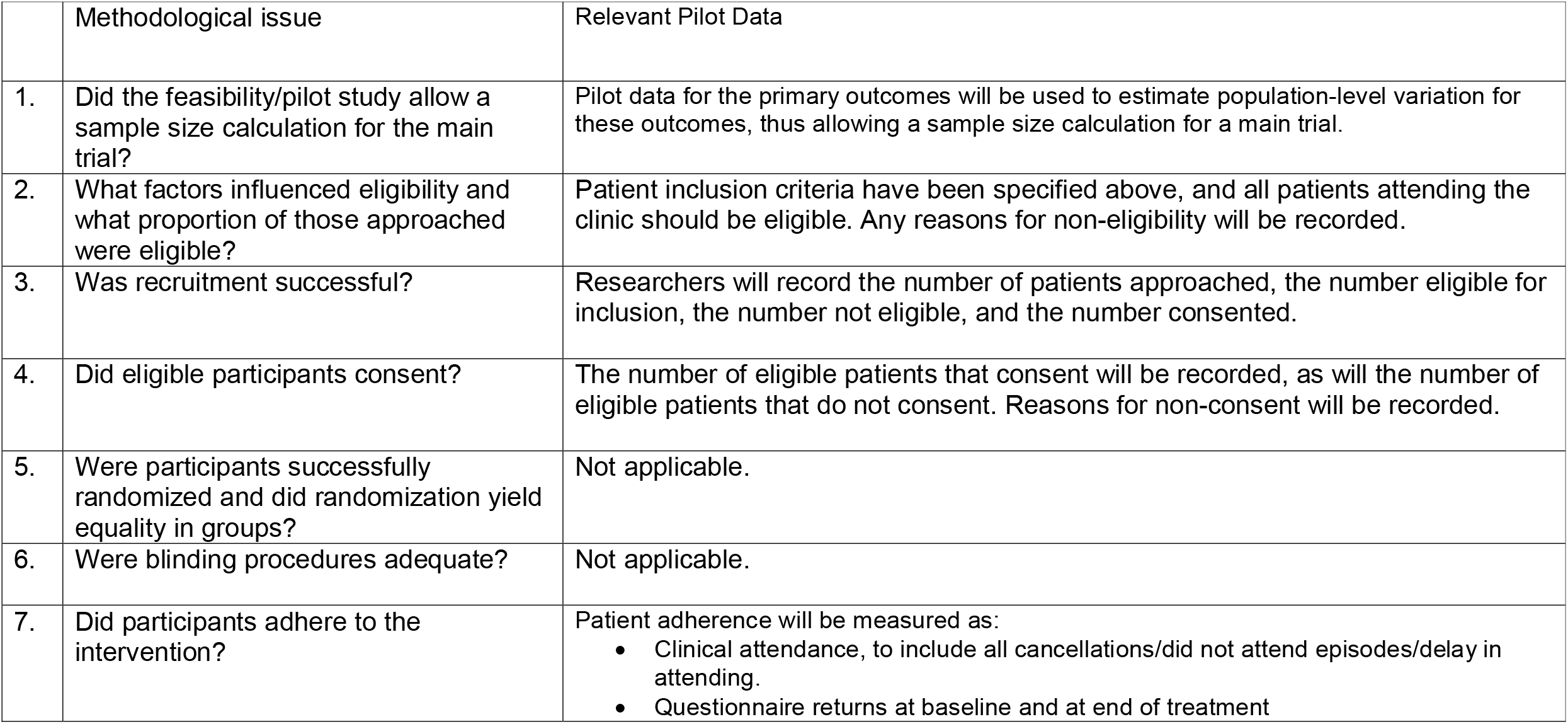
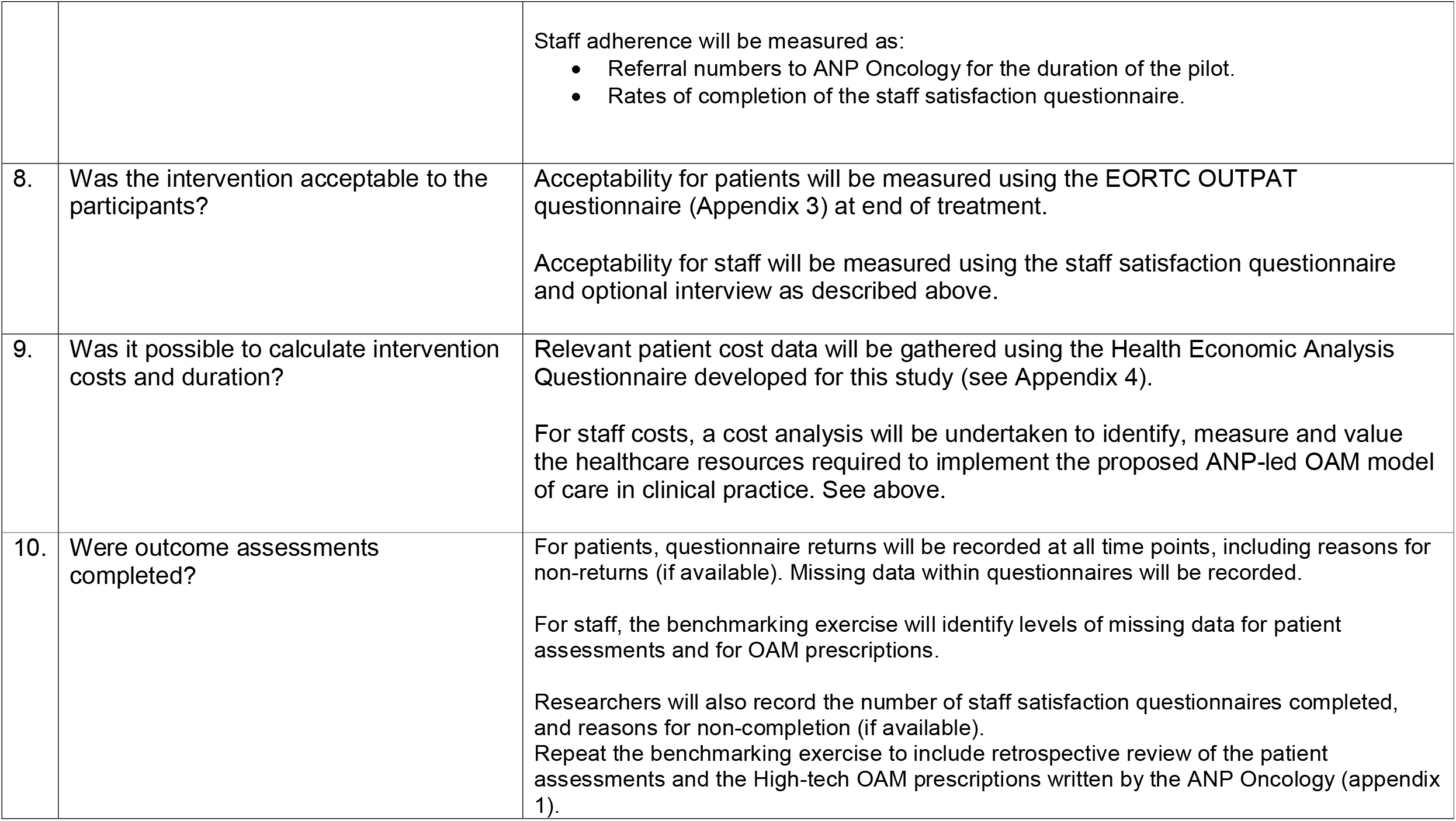
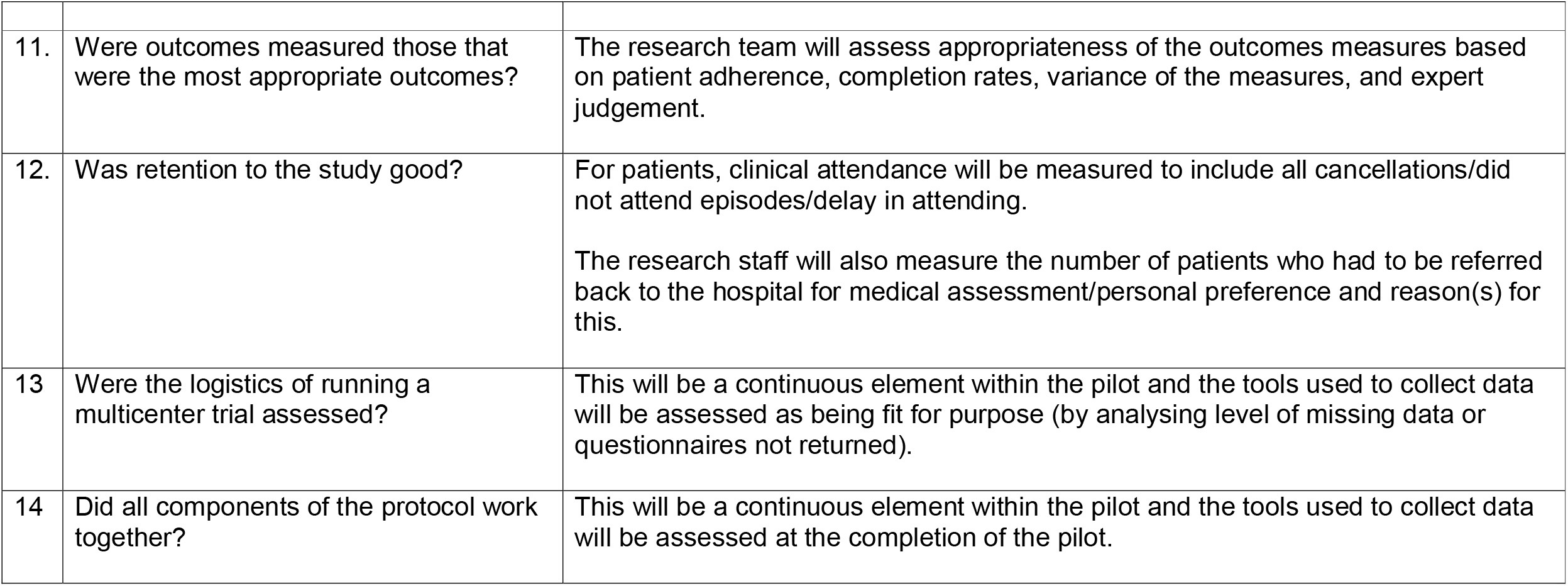
Methodological framework

|  | Methodological issue | Relevant Pilot Data |
| --- | --- | --- |
| 1. | Did the feasibility/pilot study allow a sample size calculation for the main trial? | Pilot data for the primary outcomes will be used to estimate population-level variation for these outcomes, thus allowing a sample size calculation for a main trial. |
| 2. | What factors influenced eligibility and what proportion of those approached were eligible? | Patient inclusion criteria have been specified above, and all patients attending the clinic should be eligible. Any reasons for non-eligibility will be recorded. |
| 3. | Was recruitment successful? | Researchers will record the number of patients approached, the number eligible for inclusion, the number not eligible, and the number consented. |
| 4. | Did eligible participants consent? | The number of eligible patients that consent will be recorded, as will the number of eligible patients that do not consent. Reasons for non-consent will be recorded. |
| 5. | Were participants successfully randomized and did randomization yield equality in groups? | Not applicable. |
| 6. | Were blinding procedures adequate? | Not applicable. |
| 7. | Did participants adhere to the intervention? | Patient adherence will be measured as:<br>• Clinical attendance, to include all cancellations/did not attend episodes/delay in attending.<br>• Questionnaire returns at baseline and at end of treatment |
|  |  | Staff adherence will be measured as:<br>• Referral numbers to ANP Oncology for the duration of the pilot.<br>• Rates of completion of the staff satisfaction questionnaire. |
| 8. | Was the intervention acceptable to the participants? | Acceptability for patients will be measured using the EORTC OUTPAT questionnaire (Appendix 3) at end of treatment.<br>Acceptability for staff will be measured using the staff satisfaction questionnaire and optional interview as described above. |
| 9. | Was it possible to calculate intervention costs and duration? | Relevant patient cost data will be gathered using the Health Economic Analysis Questionnaire developed for this study (see Appendix 4).<br>For staff costs, a cost analysis will be undertaken to identify, measure and value the healthcare resources required to implement the proposed ANP-led OAM model of care in clinical practice. See above. |
| 10. | Were outcome assessments completed? | For patients, questionnaire returns will be recorded at all time points, including reasons for non-returns (if available). Missing data within questionnaires will be recorded.<br>For staff, the benchmarking exercise will identify levels of missing data for patient assessments and for OAM prescriptions.<br>Researchers will also record the number of staff satisfaction questionnaires completed, and reasons for non-completion (if available). Repeat the benchmarking exercise to include retrospective review of the patient assessments and the High-tech OAM prescriptions written by the ANP Oncology (appendix 1). |
| 11. | Were outcomes measured those that were the most appropriate outcomes? | The research team will assess appropriateness of the outcomes measures based on patient adherence, completion rates, variance of the measures, and expert judgement. |
| 12. | Was retention to the study good? | For patients, clinical attendance will be measured to include all cancellations/did not attend episodes/delay in attending.<br>The research staff will also measure the number of patients who had to be referred back to the hospital for medical assessment/personal preference and reason(s) for this. |
| 13 | Were the logistics of running a multicenter trial assessed? | This will be a continuous element within the pilot and the tools used to collect data will be assessed as being fit for purpose (by analysing level of missing data or questionnaires not returned). |
| 14 | Did all components of the protocol work together? | This will be a continuous element within the pilot and the tools used to collect data will be assessed at the completion of the pilot. |

### Study setting

The on-going management of patients who are established on OAMs will, for the pilot study, be performed at a medical practice in a primary care building in Letterkenny, Co. Donegal, Ireland. This venue is external to the LUH Oncology Department, and has availability for a maximum of two half-days per week to accommodate the OAM clinic. Once patients are stable on their OAM treatment (which generally is after two cycles of treatment) the Medical Oncology team formally refers the patient to the ANP oncology for on-going assessment and monitoring for the third cycle onwards. A cycle of OAM treatment is usually 3-4 weeks in duration. The approach of reviewing the patient in the hospital oncology department for their first two cycles (or more should they be unstable on treatment initially) facilitates any major toxicities to be managed within the acute hospital setting at their review and maximises patient safety. Virtual assessments can be facilitated at subsequent cycles when patients are stable on OAMs if this is clinically acceptable and preferable to the patient (see Figure 1, Appendix 1, and Appendix 5).

**Figure 1:**
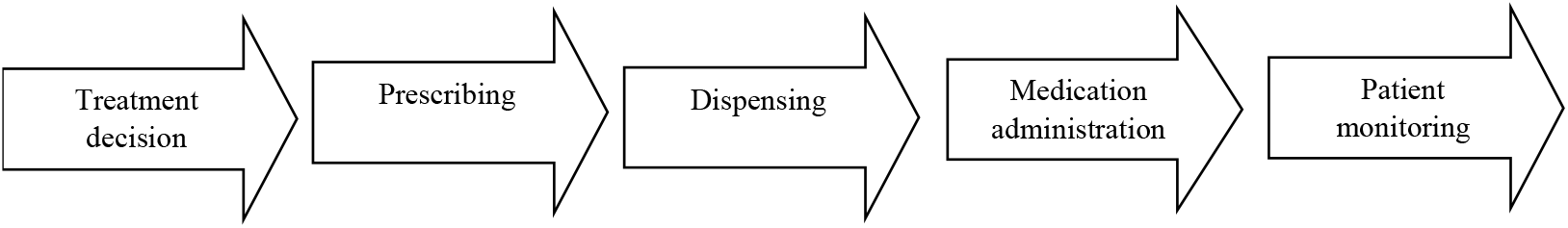
OAM processes (NCCP, 2018)

### Recruitment

All participants will be recruited from the cohort of patients being cared for by the ANP in the hospital-based OAM clinic. Patient eligibility criteria for this existing clinic are:

- Over 16 years of age.
- Have an oncology diagnosis (i.e. solid tumour).
- Be under the care of a Consultant Medical Oncologist.
- Have been prescribed OAMs

The cyclic nature of OAMs requires that patients be assessed on certain days of the week, reflective of the day they commenced treatment. This is also necessary in order to adhere to the NCCP guidelines (https://www.hse.ie/eng/services/list/5/cancer/profinfo/chemoprotocols/oral-anti-cancer-medicines). Patients whose assessments are currently fall on the two half-days per week that the study setting is available will be approached to participate in-person by the ANP Oncology. They will be provided with a participant information sheet, which will be explained. Furthermore, they will be advised that should they not wish to participate, their care will continue with the ANP oncology and their review will be facilitated on designated assessment day in the usual hospital setting. If they are content to proceed, they will be given a consent form to read, sign at home and return in a stamped addressed envelope to another member of the research team. There are no specific days for certain treatments or patient groups to attend the ANP Oncology in the hospital-based clinic, therefore the sample enrolled in this pilot study should represent a cross-section of the entire OAM patient population within the locality.

### Primary Outcomes

The primary outcomes of the pilot will be: patient safety, patient satisfaction and the cost of an ANP-led OAM clinic in a community setting. A secondary outcome will be staff satisfaction with this new clinic.

### Sample size

The pilot duration will be 4 months (January to April 2022), a timeframe adequate to include those who are receiving OAMs on an 8 or 12 weekly assessment schedule. Considering the availability of the study setting for the pilot, approximately 7-8 patient visits to the ANP will take place weekly which equates to about one third of the entire OAM weekly patient workload. Over the period of the 4-month pilot, this equates to over 100 virtual and/or face-to-face visits.

### Data Collection

#### Patient Safety

Patient safety in an ANP-led OAM clinic will be measured twofold. Firstly, there will be a repetition of, and comparison with, the benchmarking exercise for patient assessments & OAM prescriptions, which was initially performed in Phase 1 of this study (Figure 1 and Appendix 1). This consists of a Microsoft Excel tool with 82 items with the options of yes/no/not applicable. It will be completed by one of the authors (MGK or AJ) performing a retrospective chart review of 20 patient visits which includes 20 OAM prescriptions written by the ANP. This data analysis and comparison to Phase 1 will be carried out locally by the LUH staff (AJ).

Secondly, a Microsoft® Excel® spreadsheet (Appendix 2) has been developed to capture data on the patient safety aspects including patient attendance, waiting times, timeliness of review, drug toxicities requiring medical review +/- admission, ad hoc queries from General Practitioner (GP)/community pharmacists or from patient/family since last review and clinical incidents/near misses/adverse events. This data will be captured retrospectively from a review of the patient records.

#### Patient Satisfaction

Patient satisfaction with the ANP-led OAM clinic will be measured using the 7-item EORTC-OUT-PATSAT7 instrument (Bredart et al, 2018), developed to capture the perceptions of patients with cancer regarding the service and organisation of care they receive. Ratings can range from ‘fair’ to ‘excellent’, providing scores ranging from 7 to 35, with a higher score indicating higher satisfaction (Appendix 3).

#### Cost

The economic analysis will consist of two components. Firstly, the economic burden of illness falling on patients will be estimated using the EuroQol EQ-5D-5L survey instrument, to capture health-related quality of life (Balestroni & Bertolotti, 2012 ; Schwenkglenks & Matter-Walstra, 2016) and a Health Economic Analysis questionnaire created specifically for this study (Appendix 4). This questionnaire includes a series of questions that will capture healthcare service usage, out-of-pocket expenses, and employment or education participation related impacts on 10 participants (Herdman et al, 2011). Development and preliminary testing of the new five-level version of EQ-5D (EQ-5D-5L). Quality of life research, 20(10), 1727-1736. This questionnaire will be administered to individuals at the end of pilot OR at the patient’s last visit using an allocated unique identification number (UID) to ensure participant confidentiality.

Secondly, a cost analysis will be undertaken to identify, measure and value the healthcare resources required to implement the proposed ANP-led OAM model of care in clinical practice. Resource items required for the OAM clinic will be identified by the study team. This process will be directly informed by the process flow diagram, which was developed, presented and endorsed by the advisory panel of local and national experts at the end of Phase 1 of this study (Appendix 5). Unit cost data will be identified and applied to calculate individual resource costs and the total cost per patient of implementing the proposed ANP-led OAM model of care will be estimated. This data will be extrapolated to present estimates of the healthcare budget impact to implementing the proposed model of care at local, regional and national levels.

A brief clinical workflow analysis will be carried out to identify the time required for certain aspects of care via self-reported activity tracking (Lopetegui et al, 2014).. The ANP Oncology will record the time to complete toxicity assessment, physical examination and care plan (to include OAM prescription writing). To minimise onerous timing of care in a busy clinical context, data will be collected 1 day per week for the duration of the pilot and data subsequently extrapolated from this.

#### Staff Satisfaction

Staff satisfaction will be measured through an anonymised online questionnaire with a single item: “How acceptable is the intervention to you”. Responses will be measured with a 5-point Likert scale ranging from ‘totally unacceptable’ to ‘perfectly acceptable’. Space will be given for “any further comments” and contact details of a non-clinical research team member will be provided should respondents want to arrange a confidential telephone interview to provide further feedback or comment. The staff satisfaction questionnaire link will be administered to all nursing, medical, pharmacy and administrative staff who currently work in the hospital Day Unit. This questionnaire will be administered in Month 4 of the 4-month pilot.

### Data Analysis

Methodological issues will be the central focus of this pilot, using the list of 14 methodological issues that need to be examined in feasibility research (Shanyinde et al, 2011). How each of these questions will be specifically addressed in this pilot is outlined in table 2. To direct a future definitive trial, possible solutions to identified feasibility issues will be generated using a process for decision-making after pilot and feasibility trials (ADePT) (Bugge et al, 2013).

### Data Management and Protection

All collected data will be treated confidentially in line with the seven key principles related to the processing of personal data General Data Protection Regulation requirements (Health Service Executive, 2019). All consented patients and staff will be allocated a UID and this will be used on all study data collection forms. Their identification will be recorded on a password-protected document saved on a server accessed only by one of the members of the research team (JR).

At the end of the pilot, data from the questionnaires will be posted using registered mail to NUIG staff to be inputted to SPSS for statistical analysis. Similarly, at the end of the pilot, the Microsoft Excel data will be emailed to this same research team member for eventual uploading and analysis. Using the UID on all patient data obtained adheres the overriding principle of General Data Protection regulation (GDPR) in accordance to ethical approval obtained. All of the hard copy questionnaires will then be shredded by NUIG staff.

### Study Status

At the time of submission of this study protocol, (14.02.2022) recruitment of patients has begun and consents are being obtained.

## Discussion

Development of a protocol is an essential component in the process of performing a pilot trial and provides a detailed method for data collection that can be adhered to throughout. The objectives of this pilot study will be different from those of the future definitive study as it is not designed to consider the effectiveness of the intervention but rather determine whether a definitive trial can be done (Lancaster, 2015). Piloting of a new intervention for use in a definitive trial ensures that the methodological approach taken in any future research is robust and feasible and is an important step of the development process (Lancaster, 2015). Using the ADePT framework to guide decision-making (Bugge et al, 2013), completion of this pilot will evaluate whether a definitive future trial is feasible.

## Supporting information

Richmond et al-excel data collection tool appendix 2

## Data Availability

All data produced in the present study are available upon reasonable request to the authors

## Declarations

Ethical approval has been granted by Letterkenny University Hospital’s Research Ethics Committee (17/11/2021). Informed consent was obtained before recruiting participants.

## Authors’ contributions

J.R. Supervision, conceptualisation, methodology, project administration, writing— review & editing; M.G.K conceptualisation, methodology, writing—review & editing; A.J. methodology, writing—review & editing; P.J.M. conceptualisation, methodology, review & editing; A.J. methodology, writing—review & editing; P.G. health economics methodology, review & editing, A.W.M. conceptualisation, methodology, review & editing.

## Appendix 1 Best-practice standard used for bench-marking

Presented in the logical sequence of the Systemic Anti-Cancer Treatment Model of Care identified by NCCP [2018] {Figure 1}

### 1: Treatment decision

#### Standard 1.1

Patients who are to commence on an Oral Anti-Cancer Medicine have an assessment to determine their suitability for home management of treatment (NCCP (2018).

##### Features of a service meeting this standard should include

1.1.1 Baseline documented assessment of the patient regarding their capabilities and understanding of OAM.

#### Standard 1.2

Written consent should be obtained from the patient initially prior to the Medical team commencing treatment with OAM (ONS, 2016; Hall, 2016).

##### Features of a service meeting this standard should include

1.2.1 Provision of pre-treatment timely and relevant information to patients regarding the proposed treatment to include reason for treatment, goal of therapy and potential side-effects.
1.2.2 MOATT© - MASCC tool (Teaching Tool for Patients Receiving Oral Agents for Cancer) used to assess and document suitability and understanding of OAM (NCCP Systemic Anti-Cancer Treatment assessment form)^1^.
1.2.3 Written consent should be obtained from the patient prior to commencing and a copy available in the patient’s notes.
1.2.4 Provision of information (1.2.1) and written consent (1.2.2) should be repeated should drug regime change.

### 2: Prescribing

#### Standard 2.1

Oral Anti-Cancer Medicines should be prescribed to the same safety standards as parenteral Systemic Anti-Cancer Therapy (NCCP (2018).

##### Features of a service meeting this standard should include

2.1.1 The first cycle of a course of an Oral Anti-Cancer Therapy should be written by a consultant or registrar based on the consultant’s written therapy plan (NCCP (2018).
2.1.2 Subsequent cycles should be written by a consultant, specialist registrar, registrar or Registered Nurse Prescriber following the consultant’s written therapy plan (NCCP (2018).
2.1.3 For any dose modification, a new prescription should be written by any of the Health Care Professionals listed in 2.1.2 (NCCP (2018).
2.1.4 Oral Anti-Cancer Medicine prescriptions should be written for one cycle only except when a patient’s scheduled review is longer than one cycle (NCCP (2018).
2.1.5 All Oral Anti-Cancer Medicine prescriptions should be verified by an oncology pharmacist (NCCP 2018).

#### Standard 2.2

OAM prescriptions should include adequate information to facilitate prescription review by the community pharmacist (NCCP, 2018).

##### Features of a service meeting this standard should include

2.2.1 An OAM prescription should include the following information (unless otherwise stated this is as per NCCP (2018))^1^:
  - Patient name
  - Patient address
  - Date of Birth
  - Hospital number
  - Patients consultant
  - Height/weight/Body Surface Area (BSA) (if relevant)
  - Diagnosis
  - Diagnosis code
  - Frequency of patient review
  - Allergies/sensitivities/contraindications
  - Protocol name
  - Protocol code
  - Deviations from protocol
  - Course number
  - Medication name (generic name but biosimilars require trade name also) (LUH^1^)
  - Medication dose (to include M2 if relevant)
    - Doses rounded to the nearest tablet size (ASCO, 2013)
    - Doses do not include trailing zeros (ASCO, 2013).
    - Use a leading zero for doses (ASCO, 2013).
  - Medication administration frequency
  - Number of treatment days
  - Route (ONS, 2016)
  - Total quantity to be dispensed (ONS, 2016).
    - Wording should read to not dispense more than x tablets/capsules (LUH).
  - Planned treatment start date and/or cycle prescription commences
  - Signature and printed name.
  - Clinician registration number
  - Date
  - Contact details of prescriber
  - GP name & address
  - Nominated pharmacy & address

### 3: Dispensing

#### Standard 3.1

To facilitate appropriate counselling of the patient, the first cycle of Oral Anti-Cancer Medicines and the first cycle of a dose adjustment should be dispensed in a hospital setting where possible (NCCP, 2018) (exception mentioned by NCCP, 2018).

##### Features of a service meeting this standard should include

3.1.1 Face-to-face assessment of the patient by an Oncology health care professional pre first treatment with OAM.
3.1.2 Face-to-face assessment of the patient by an Oncology health care professional in the event of an OAM dose adjustment.

#### Standard 3.2

Only one cycle of an OAM should be dispensed at a time (NCCP, 2018)^1^

##### Features of a service meeting this standard should include

3.2.1 The prescription should identify the number of days treatment and/or the number of medications to be dispensed (NCCP, 2018).

#### Standard 3.3

The quantity dispensed should not exceed the number of doses required to complete the cycle (NCCP, 2018)^2^.

##### Features of a service meeting this standard should include

3.3.1 Days supply limitation rules should be identified on the prescription (Battis et al, 2016).

### 4: Medication administration

#### Standard 4.1

Patients prescribed Oral Anti-Cancer Medicines should have access to standardised education to support safe administration, safe handling, and management of side effects (NCCP, 2018)

##### Features of a service meeting this standard should include

4.1.1 Dedicated initial pre-treatment face-to-face education session with an Oncology Health Care Professional (Muluneh et al, 2018) detailing storage, handling/preparation, administration, disposal of oral chemotherapy, possible drug/drug and drug/food interactions and plan for missed doses and management of side effects of OAM and documented appropriately (ASCO, 2013; NCCP, 2018).
4.1.2 MOATT© - MASCC tool (Teaching Tool for Patients Receiving Oral Agents for Cancer) used to frame, check understanding and document the pre-treatment education (NCCP Systemic Anti-Cancer Treatment assessment form)^1^.
4.1.3 Patients should be informed of the required monitoring arrangements and have access to information in the written protocol and treatment plan from the hospital where treatment was initiated (UKONS, 2010).

### 5: Patient monitoring

#### Standard 5.1

Patients should not commence their treatment until the results of their monitoring tests are known (NCCP 2018)

##### Features of a service meeting this standard should include

5.1.1 Documented review of monitoring tests and contact with the patient prior to commencing a treatment cycle (NCCP, 2018).

#### Standard 5.2

Patients on Oral Anti-Cancer Medicine treatment should be reviewed by specialist health care professional in an appropriate location at the predefined intervals (NCCP, 2018) prior to every treatment cycle (UKONS, 2010).

##### Features of a service meeting this standard should include

5.2.1 Assessment of the patient^1^ by an Oncology health care professional in an appropriate location at the predefined intervals (NCCP, 2018) as identified by the NCCP OAM protocols^2^ prior to every treatment cycle (UKONS, 2010).
  5.2.2 Completion of patient assessment template specific to an OAM can standardise medication monitoring and facilitate documentation of patient interactions and interventions (Battis et al, 2016).
  5.2.3 Completion of patient telephone assessments in a template which allows flexibility in the conduction of the phone follow up and can be performed by a specialist oncology health care professional team member (May et al, 2017).
5.2.4 Patient assessment requires determining adherence (Muluneh et al, 2018)^3^.
5.2.2 Patient assessment includes but not limited to documentation of the following (laboratory tests required is as per NCCP protocol for specific regime) (unless otherwise stated this is per NCCP SACT assessment forms)^4^:
  - Primary diagnosis
  - Allergies^1^
  - Frequency of review
  - NCCP regime
  - Cycle number
  - Day number
  - Translator present (changed to translator required)
  - Has the patient been admitted to hospital or seen their GP since their last treatment?
    ▪ Did the patient receive a discharge prescription?
  - Any patient infection control alert/issues?
  - Grading of toxicities
  - Performance status documented
  - Vital Signs/Early Warning Score (EWS) completed
  - MST (Malnutrition assessment TOOL) Score completed
  - Tumour markers as per medical instruction
  - LMP (last menstrual period) documented (if applicable)
  - Urinalysis completed (if applicable)
  - MOATT© - MASCC tool completed (Teaching Tool for Patients Receiving Oral Agents for Cancer).
    ▪ Assessment of ability to manage treatment at home
  - Weight in metrics (ISMP, 2020)
  - Is there a need for a follow up phone call?
  - Any dose modifications or any cycle delays (UKONS, 2010)
  - Is there any change to medication?
  - Is there a history of an adverse event on a previous cycle?
  - Disease monitoring and any other test/s as directed by the supervising Consultant (NCCP OAM protocols^2^)
  - At defined intervals the following should be checked (specific to each drug) and reviewed as satisfactory prior to proceeding with treatment (NCCP OAM protocols).
  - HCG (pregnancy) test completed (if applicable)
    ▪ FBC, renal and liver profile
    ▪ Calcium level (JR)
    ▪ Serum glucose
    ▪ Blood pressure
    ▪ Urinalysis
    ▪ Thyroid function tests
    ▪ ECG if clinically indicated or if history of cardiac problems
    ▪ Total cholesterol and triglycerides
    ▪ Other ________________

#### Standard 5.3

Patients to have contact post commencement of cycle 1 of treatment.

##### Features of a service meeting this standard should include

5.3.1 Follow up phone call by pharmacist to patient after 72 hours of commencing cycle 1 (Calabrese et al, 2015).
5.3.2 High risk patients to have a phone call made to them days 8 and 10 (Deluche et al, 2020).
5.3.3 All patients to have a phone call made to them days 15 & 30 (Deluche et al, 2020).
  - Contact or visits are beneficial especially in the 1st 2 cycles to improve treatment and symptoms experience (Vidall, 2010^1^; Oakley et al, 2014; Wong et al, 2016).

#### Standard 5.4

Clinical SACT services should provide a 24-hour telephone advice service for patients prescribed oral therapies, with appropriately trained nursing, medical or pharmacy staff handling queries (UKONS, 2010; Deluche et al, 2020).

##### Features of a service meeting this standard should include

5.4.1 All patients/families to be provided with 24hr/7 day contact details of the hospital at commencement of cycle 1 of OAM.
5.4.2 All patients should have access to advice from an appropriately qualified healthcare professional with experience in cancer treatment in the hospital (UKONS 2010).

#### Standard 5.5

Information strategies should be developed to standardise communication between hospitals, community pharmacists and GPs (NCCP, 2018) and primary care nurses and GPs require precise and regular information (UKONS, 2010).

##### Features of a service meeting this standard should include

5.5.1 Adherence to standard 2.2 will facilitate this with community pharmacists.
5.5.2 Letter to GP to be sent if patient condition changes, medication changes or the dose changes. If there is stable care for a period of time beyond 6 months then a GP letter should be sent to provide an update (JR).
5.5.3 After the final cycle of a treatment course, the records for each patient should include the following (UKONS, 2010) and communicated to GP (JR):
  - Whether the course was completed or not.
  - If the course was not completed or if the planned dose was reduced, the reasons for cessation or reduction.
  - For completed courses of non-adjuvant treatment, a reference to the response.
  - The plans for on-going review and support.

## Appendix 2 Microsoft® Excel® Data collection tool (using UID)

Excel Tab 1: Data collected (demographics):

- Gender.
- Age.
- Primary cancer.
- OAM specific regime/medication.
- OAM cycle (for first assessment on pilot).
- Aim of treatment (i.e. curative/palliative).
- Date of cancer diagnosis.
- Date of most recent recurrence.

Excel Tab 2 Data collected (geographical):

- Distance (in kilometres) travelled for OAM assessment.
- Methods of transport (i.e. car/bus).
  - Own or other driver (if car used).
  - Time off work for other driver (if has to be driven)-measured in minutes for round trip.
- Amount spent in parking (if car used).
- Amount spent for travel (if public transport used).
- Closest primary care centre to patient’s home (measured in kilometres)
- Closest community hospital to patient’s home (measured in kilometres)

Excel Tab 3 Data collected (appointment details):

- Type of assessment (i.e. physical assessment or virtual assessment).
- Did they attend on time (i.e. cancel/did not attend/attended on time).
  - Extent of time in delay (measured in minutes from appointment time) if relevant.
  - Reason for delay in attending if relevant.
- Patient reviewed at correct interval as per protocol.
  - Extent of interval delay (measured in days) if relevant.
  - Reason for any interval delay.
- Waiting time to be reviewed by ANP Oncology (measured in minutes from appointment time).
- Total time spent with ANP (measured in minutes)
  - Specific time (measured in minutes) performing observations, measuring weight and performing toxicity assessment (could be done by other nursing grades).
  - Specific time (measured in minutes) communicating outcome of assessment to General Practitioner and/or community pharmacist.
- Outcome of review (i.e. proceed with OAM/hold OAM/discontinue OAM).
- Toxicities requiring referral to hospital for medical review.
  - Description of toxicity (if relevant).
- Toxicities/problem requiring admission
  - Description of toxicity/problem if relevant.
- Adverse events previous cycle.
  - Description of adverse events if relevant
- Near misses previous cycle.
  - Description of near misses if relevant.
- Clinical incident previous cycle.
  - Description clinical incident if relevant.
- Number of ad hoc queries from General Practitioner in previous cycle.
- Number of ad hoc queries from Community pharmacist in previous cycle.
- Number of ad hoc queries from other HCP in previous cycle.
- Number of ad hoc queries from patient in previous cycle.
- Number of ad hoc queries from family in previous cycle.

## Appendix 3 EORTC OUT-PATSAT7

© QLQ-OUT-PATSAT7 Copyright 2017 EORTC Quality of Life Group. All rights reserved.

### YOUR EXPERIENCE OF OUTPATIENT CARE

#### EORTC OUT-PATSAT7

We are interested in your MOST RECENT experience of the care received in the outpatient setting in this clinic. Please answer all the questions yourself by circling the number that best applies to you. There are no ‘right’ or ‘wrong’ answers

**In the outpatient setting in this hospital, how would you rate <u>services and care organisation</u>, in terms of:**

|  | Poor | Fair | Good | Very<br>good | Excellent |
| --- | --- | --- | --- | --- | --- |
| 34. The opportunity to see the same caregivers<br>when you come to the outpatient clinic? | 1 | 2 | 3 | 4 | 5 |
| 35. The ease of arranging medical appointments<br>at convenient times? | 1 | 2 | 3 | 4 | 5 |
| 36. The waiting time before obtaining<br>a medical appointment? | 1 | 2 | 3 | 4 | 5 |
| 37. The ease of communicating with the hospital<br>services from home? | 1 | 2 | 3 | 4 | 5 |
| 38. The information provided about what you<br>should/should not do after you leave your<br>hospital appointment? | 1 | 2 | 3 | 4 | 5 |
| 39. The information on who to contact if you are<br>worried after you leave your hospital<br>appointment? | 1 | 2 | 3 | 4 | 5 |
| 40. The provision of follow-up by the different<br>caregivers (doctors, nurses, physiotherapists,<br>psychologists, etc.) after treatment? | 1 | 2 | 3 | 4 | 5 |

## Appendix 4 Health Economic Analysis Questionnaires

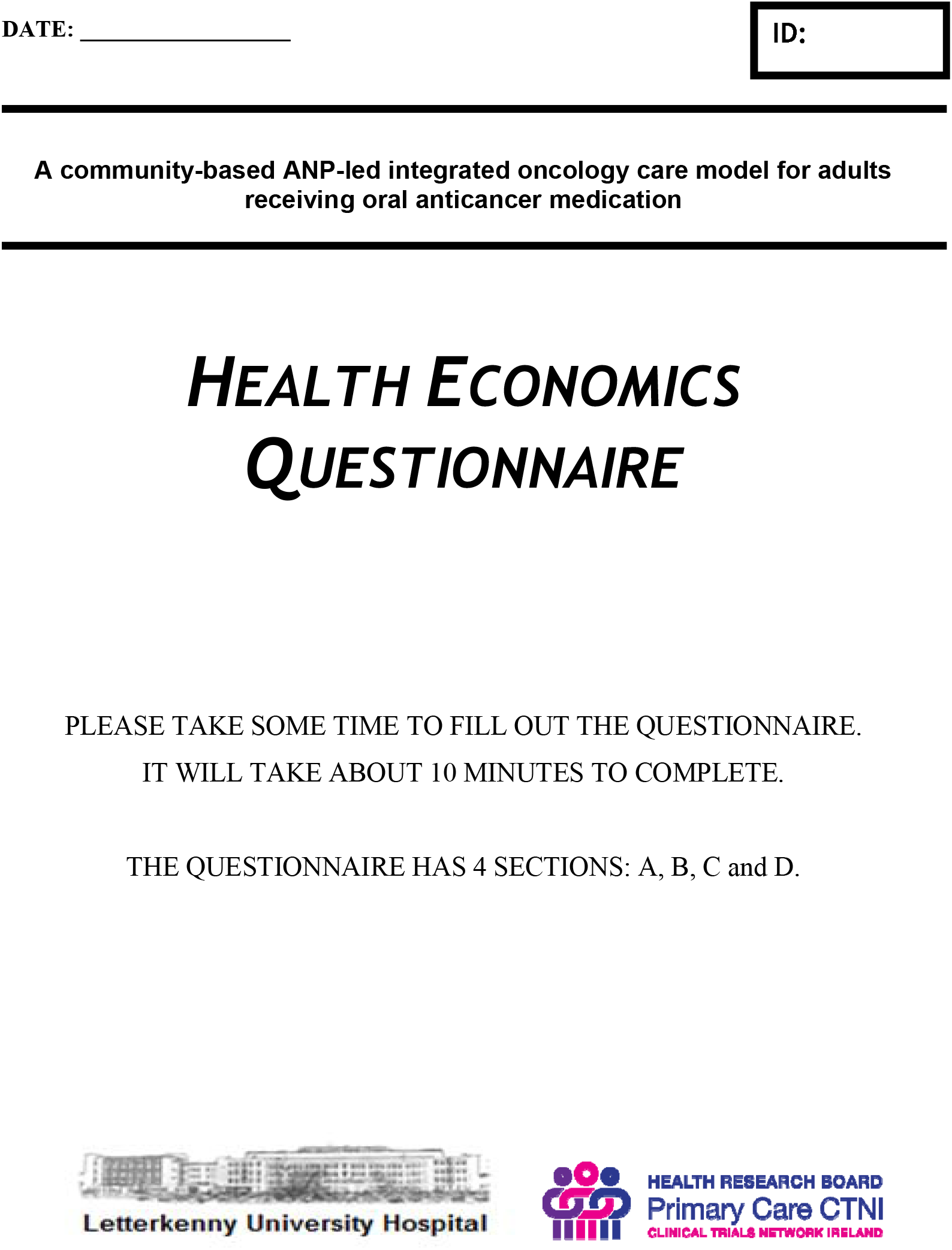

In this questionnaire we are trying to find out how **you are feeling** and about the **costs** to you arising from your cancer diagnosis and your current treatment. **Your answers are important** because they will give people in the Health Service an idea of how much the treatment costs you. Apart from the researchers, other nurses and doctors and health care professionals involved in your care **will not have access** any of the information you provide.

The information you give will be **completely confidential** and you will **not be identified** in any way. The detail you provide will be **held securely** as protected information.

### Please try and answer every question

If you are not sure or cannot remember the exact details, please give the best answer you can. If you **do not want to answer a question** for whatever reason, then just leave it blank.

#### THANK YOU VERY MUCH FOR YOUR TIME AND HELP

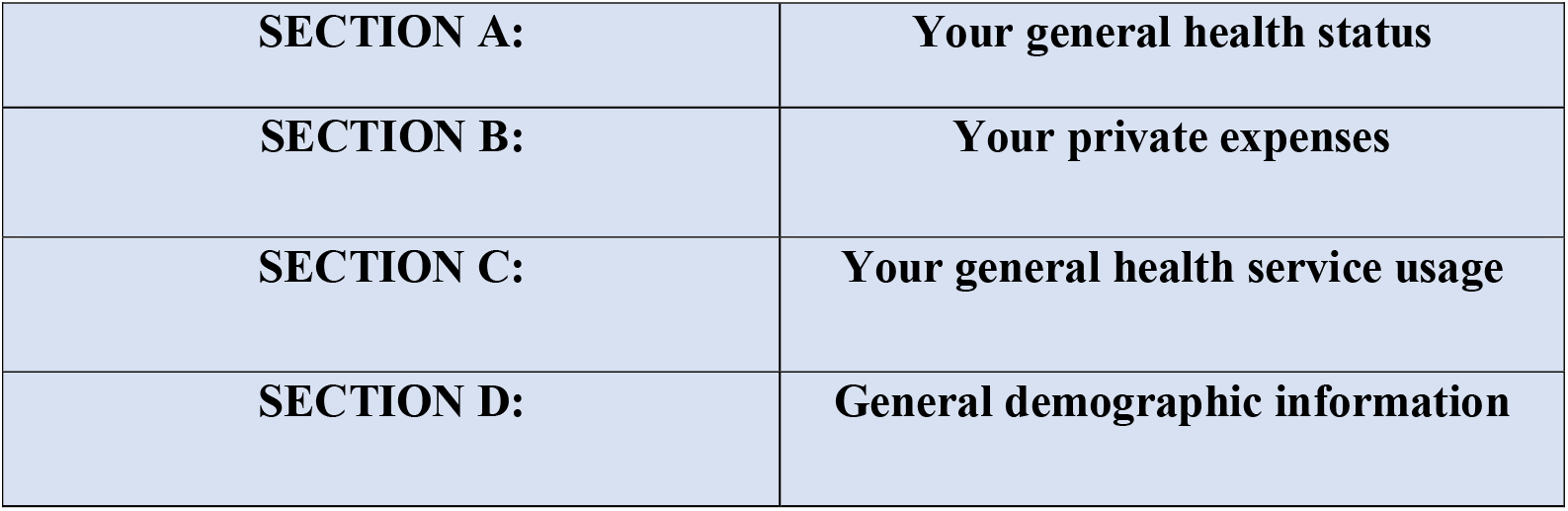

### SECTION A: YOUR GENERAL HEALTH STATUS

We are interested in how you feel about **your health today**.

#### EQ-5D-5L

**Under each heading, please tick the ONE box that best describes your health TODAY**

**Mobility**

I have no problems in walking about
I have slight problems in walking about
I have moderate problems in walking about
I have severe problems in walking about
I am unable to walk about

**SELF-CARE**

I have no problems washing or dressing myself
I have slight problems washing or dressing myself
I have moderate problems washing or dressing myself
I have severe problems washing or dressing myself
I am unable to wash or dress myself

**Usual Activities (e.g**., **work, study, housework, family or leisure activities)**

I have no problems doing my usual activities
I have slight problems doing my usual activities
I have moderate problems doing my usual activities
I have severe problems doing my usual activities
I am unable to do my usual activities

**Pain/Discomfort**

I have no pain or discomfort
I have slight pain or discomfort
I have moderate pain or discomfort
I have severe pain or discomfort
I have extreme pain or discomfort

**Anxiety/Depression**

I am not anxious or depressed
I am slightly anxious or depressed
I am moderately anxious or depressed
I am severely anxious or depressed
I am extremely anxious or depressed

© EuroQol Research Foundation. EQ-5D™ is a trade mark of the EuraQol Research Foundation

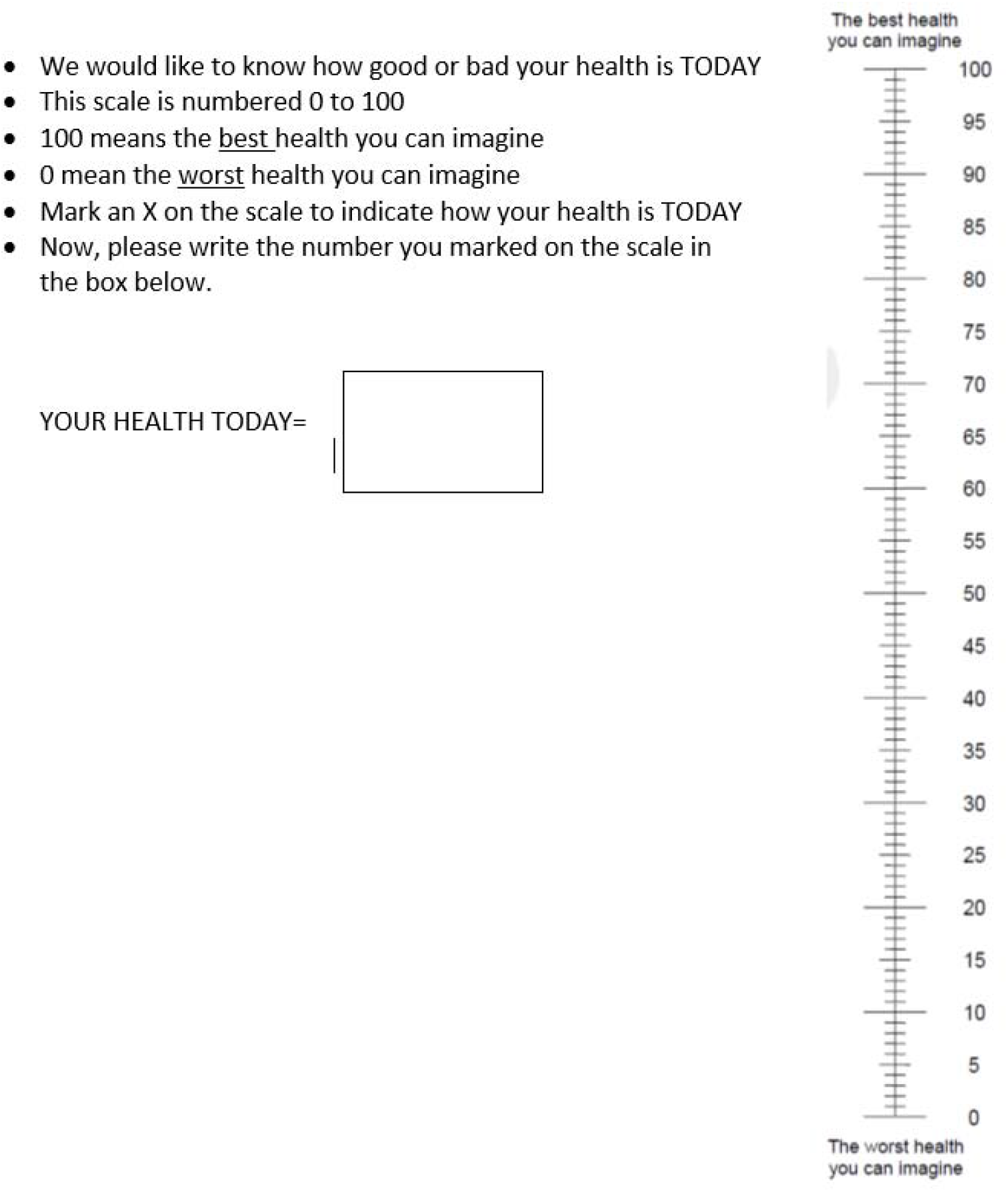

### SECTION B: YOUR PRIVATE EXPENSES

#### This information gives the research team an understanding of the financial costs of having cancer and needing cancer treatment

**Q1**. We are interested in whether you have missed **work** or arrived late/left early to attend the oncology clinic appointment with the ANP in the **last 3 months. If you have**, please fill in the section below and if not then move onto question 2.

| TYPE |  | FREQUENCY |
| --- | --- | --- |
| **YOUR TIME in the last 3 months** |  |  |
| In the **last 3 months**, did you miss **work** to attend **appointments**? | ☐ YES<br>☐ NO<br>☐ Not applicable (NA) | If **yes**, how many **days** did you miss? (Please estimate) |
|  |  | If **yes**, how many **hours** did you miss? (Please estimate) |
| In the **last 3 months**, did you arrive late or leave early from **work** because you were **unwell**? | ☐ YES<br>☐ NO<br>☐ NA | If **yes**, how many **times** did this happen? (Please estimate) |
| In the **last 3 months**, did you miss **work** because you were **unwell**? | ☐ YES<br>☐ NO<br>☐ NA | If **yes**, how many **days** did you miss? (Please estimate) |

**Q2**. We would like to know what the **financial costs** are for your healthcare. Please tick which of the following you have to **pay for** and give us an estimate of how much you have had to pay over the last **3 months. If you have costs**, please fill in the section below and if not then move onto question 3.

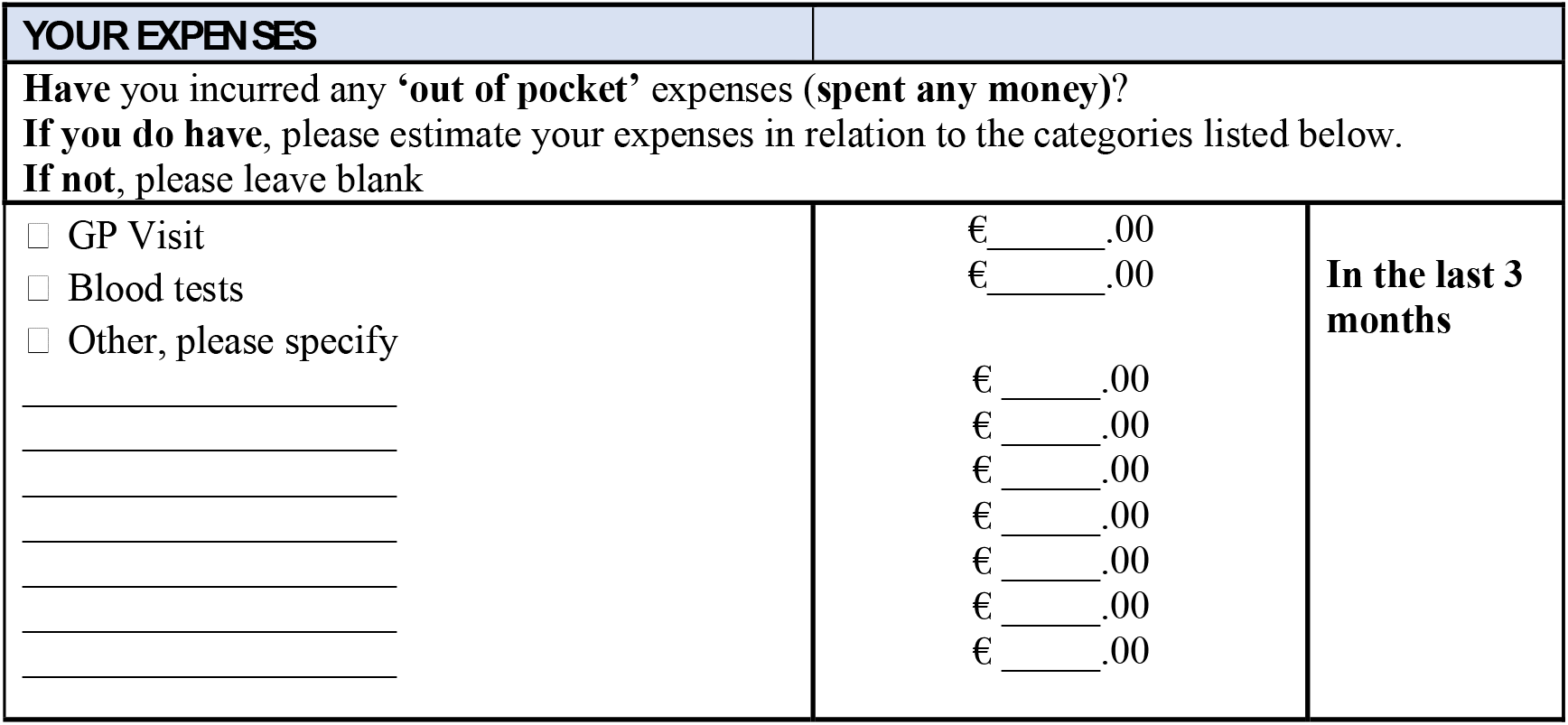

**Q3**. We are interested in whether **you spend any money** attending your appointments with the ANP for your oral anti-cancer treatment. **If you do**, please estimate how much you pay **per appointment**. If you don’t have any costs, then move on to question 4.

| YOUR CARE COSTS | Attending for blood tests | Attending the ANP oncology clinic |
| --- | --- | --- |
| Do you usually get someone to look after your children when you have an appointment? | ☐ YES ☐ NO ☐ NA | ☐ YES ☐ NO ☐ NA |
| If yes, how much do you usually pay them per appointment? | €_____.00 |  |
| Do you usually get someone to look after other relatives when you have an appointment? | ☐ YES ☐ NO ☐ NA | ☐ YES ☐ NO ☐ NA |
| If yes, how much do you usually pay them per appointment? | €_____.00 |  |

**Q4**. We would like to know if there are any **travel expenses** for you to attend your **appointments** (e.g., petrol, bus fare etc.). **If you do**, please estimate how much **per appointment**.

| YOUR TRAVEL COSTS | For blood tests | Attending the ANP oncology clinic |
| --- | --- | --- |
| **How many kilometres (km)** do you have to travel from your home to your appointment? | _____ km | _____ km |
| **How** do you usually travel to these appointments?<br>(e.g., **bus, car, train, taxi, walk, other**)<br>Please list **all** that apply. |  |  |
| If you drive how much do you usually spend on **parking** per appointment? | €_____.00 | €_____.00 |
| If you use **public transport** how much do you usually spend per appointment? | €_____.00 | €_____.00 |
| If you usually **pay someone** to take you to your appointment how much do you usually spend per appointment? | €_____.00 | €_____.00 |

### SECTION C: YOUR GENERAL HEALTH SERVICE USAGE

**Q1**. We are interested in whether you have used any of the **<u>healthcare services</u>** listed below in the last **3 months**. If you have used a service, please indicate how **many visits** or **how often** you used that service in the last **3 months**.

| TYPE | FREQUENCY OF USE |  |
| --- | --- | --- |
| SERVICE | Have you used this service in the last **3 months**? | If **YES**, approximately how **many visits** or **how often** did you use this service? |
| **Outpatient clinic** | ☐ YES ☐ NO →<br>(Please tick) | **Visits:** Number____<br>**Phone calls:** Number____ |
| **Dietician Services** | ☐ YES ☐ NO →<br>(Please tick) | **Visits:** Number____<br>**Phone calls:** Number____ |
| **Optician Services** | ☐ YES ☐ NO →<br>(Please tick) | **Visits:** Number____<br>**Phone calls:** Number____ |
| **Audiology (hearing) service** | ☐ YES ☐ NO →<br>(Please tick) | **Visits:** Number____<br>**Phone calls:** Number____ |
| **General Practitioner (GP)** | ☐ YES ☐ NO →<br>(Please tick) | **Visits:** Number____<br>**Phone calls:** Number____ |
| **Practice Nurse (In GP practice)** | ☐ YES ☐ NO →<br>(Please tick) | **Visits:** Number____<br>**Phone calls:** Number____ |
| **Public Health Nurse** | ☐ YES ☐ NO →<br>(Please tick) | **Visits:** Number____<br>**Phone calls:** Number____ |
| **Physiotherapy Services** | ☐ YES ☐ NO →<br>(Please tick) | **Visits:** Number____<br>**Phone calls:** Number____ |
| **Occupational Therapist** | ☐ YES ☐ NO →<br>(Please tick) | **Visits:** Number____<br>**Phone calls:** Number____ |
| **Social Worker** | ☐ YES ☐ NO →<br>(Please tick) | **Visits:** Number____<br>**Phone calls:** Number____ |
| **Psychological or Counselling Services** | ☐ YES ☐ NO →<br>(Please tick) | **Visits:** Number____<br>**Phone calls:** Number____ |
| **Other** | ☐ YES ☐ NO →<br>(Please tick and identify the service) | _____<br>_____<br>_____ |

### SECTION D: YOUR DEMOGRAPHICS

**We are interested in learning more about you. This information is helpful to the research team but is not essential for you to complete**.

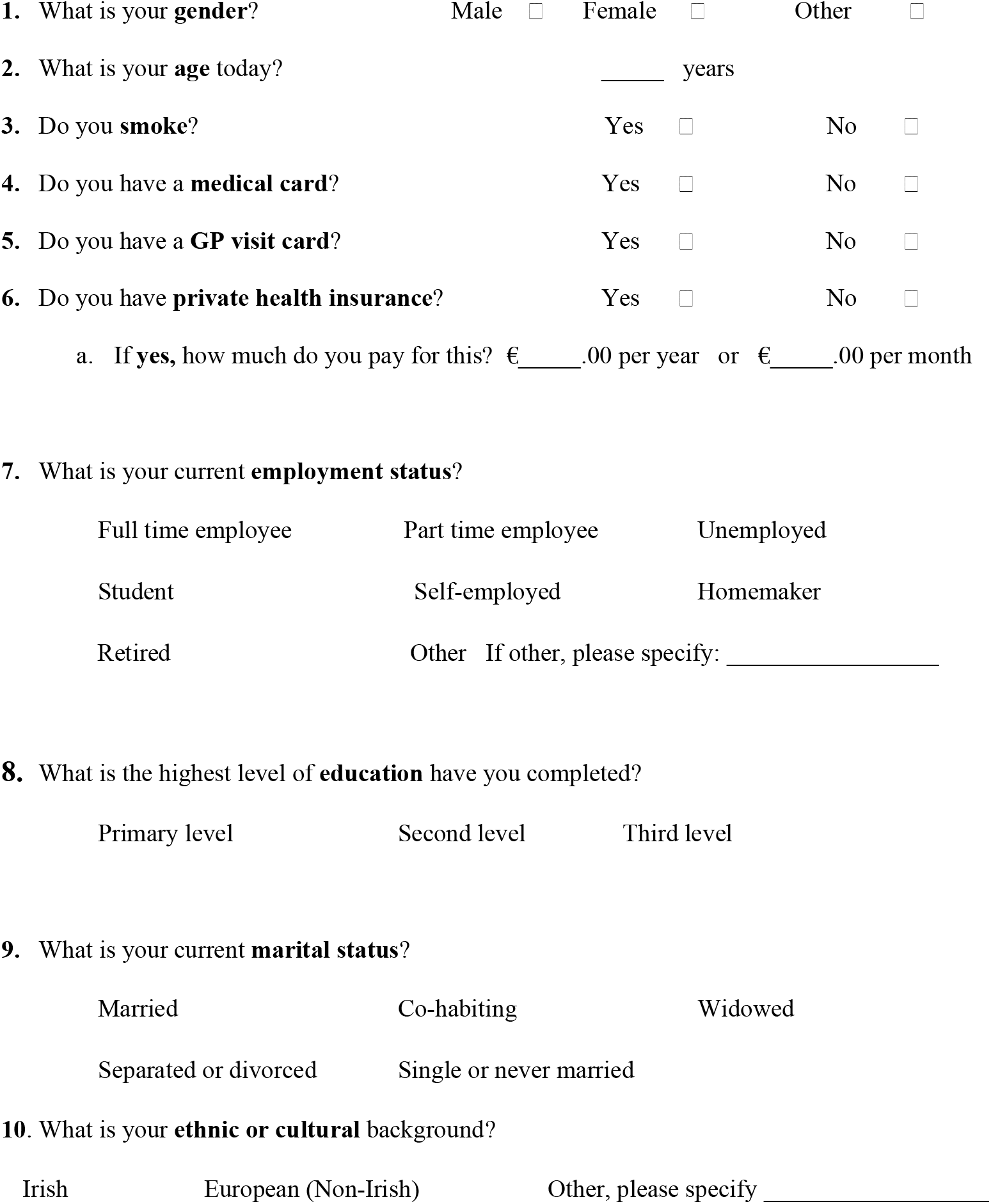

|  |  |  |
| --- | --- | --- |
| Full time employee | Part time employee | Unemployed |
| Student | Self-employed | Homemaker |
| Retired | Other If other, please specify: _____ |  |
| 8. What is the highest level of **education** have you completed? |  |  |
| Primary level | Second level | Third level |
| 9. What is your current **marital status**? |  |  |
| Married | Co-habiting | Widowed |
| Separated or divorced | Single or never married |  |
| 10. What is your **ethnic or cultural** background? |  |  |
| Irish | European (Non-Irish) | Other, please specify _____ |

If you would like to provide feedback as to how you felt answering these questions, please do so:

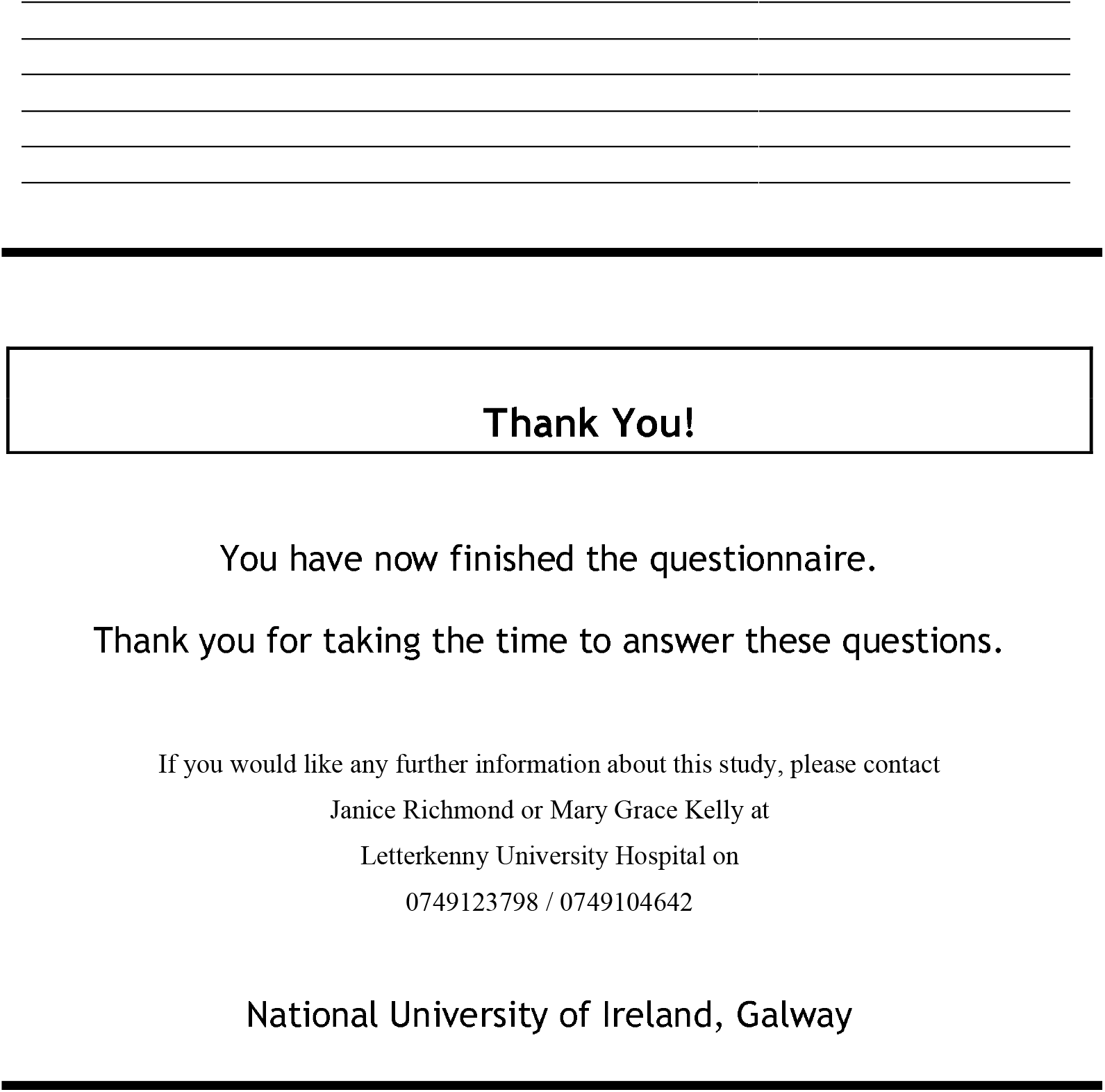

## Appendix 5 Proposed Model for OAM Care (Pilot)

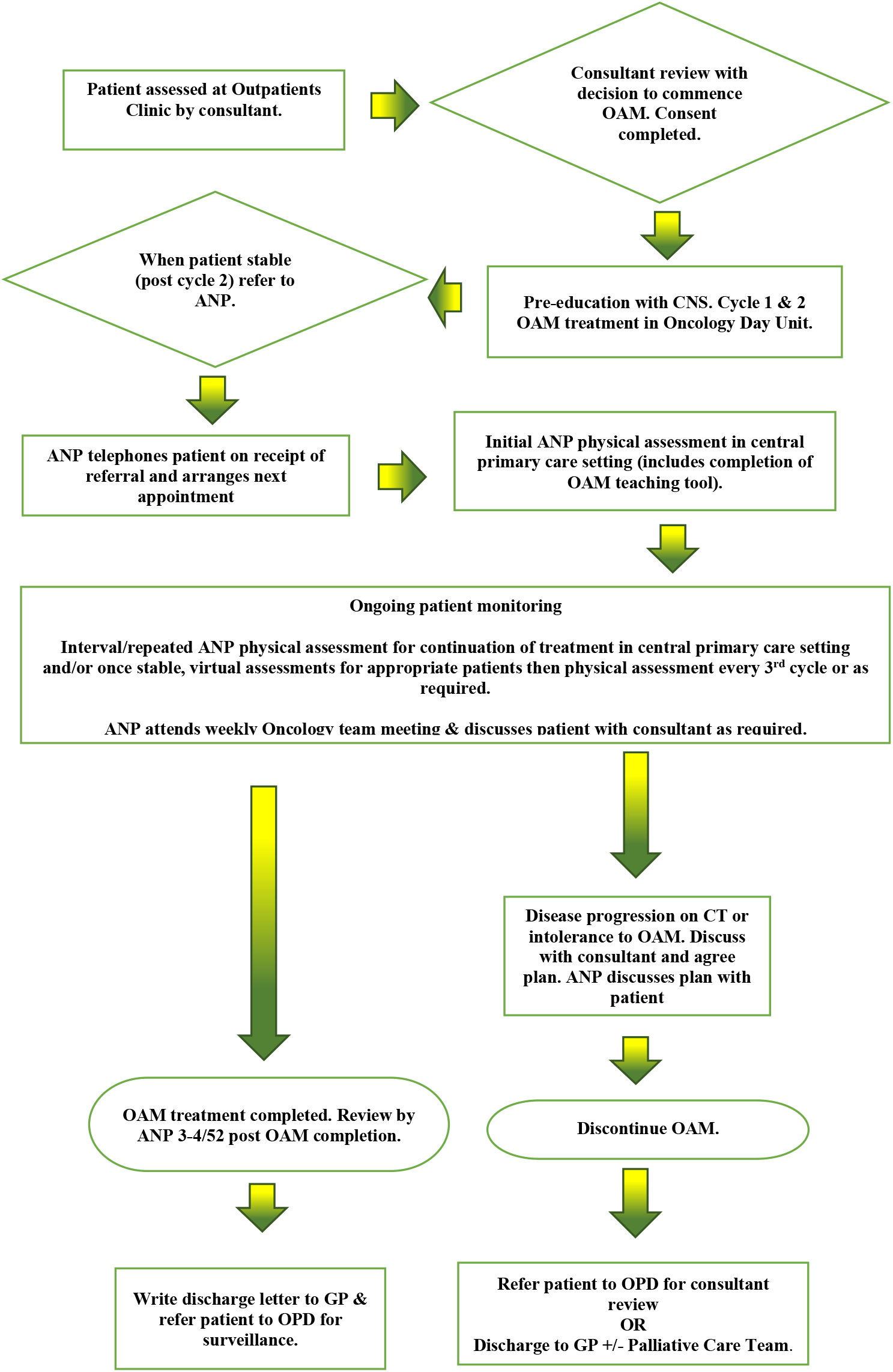

## Footnotes

1 https://www.hse.ie/eng/services/list/5/cancer/profinfo/medonc/sactguidance/extravasation.html

1 Email orders are considered written orders (ASCO, 2013).

1 LUH is Letterkenny University Hospital (author’s place of work)

1 Community pharmacy dispenses the medication so the treating hospital team have no control over this aspect of care.

2 Community pharmacy dispenses the medication so the treating hospital team have no control over this aspect of care.

1 https://www.hse.ie/eng/services/list/5/cancer/profinfo/medonc/sactguidance/extravasation.html

1 Monitoring laboratory tests is not sufficient for patient management pts on OAMs. Close monitoring and follow up of patients on OAMs is crucial to achieve intended therapeutic outcome, improve drug safety and adherence and to reduce drug’s adverse events and healthcare cost (Battis et al, 2016)

2 https://www.hse.ie/eng/services/list/5/cancer/profinfo/chemoprotocols/oral-anti-cancer-medicines/

3 No benefit noted to treatment calendars (Wong et al, 2014)

4 https://www.hse.ie/eng/services/list/5/cancer/profinfo/medonc/sactguidance/extravasation.html

1 Allergies should be checked prior to prescribing and not required at each review (JR)

2 https://www.hse.ie/eng/services/list/5/cancer/profinfo/chemoprotocols/oral-anti-cancer-medicines/

1 Vidall (2010) used home visits to monitor patients

